# COVID treatment and in-hospital length of stay inequalities between race in the US over time

**DOI:** 10.1101/2022.09.30.22280586

**Authors:** Benjamin M. Althouse, Charlotte Baker, Peter D Smits, Samuel Gratzl, Ryan H Lee, Brianna M Goodwin Cartwright, Michael Simonov, Michael D Wang, Nicholas L Stucky

## Abstract

**Introduction:** Demonstrated health inequalities persist in the United States. SARS-CoV-2 (COVID) has been no exception, with access to treatment and hospitalization differing across race or ethnic group. Here we aim to assess differences in treatment with remdesivir and hospital length of stay across four waves of the pandemic.

**Methods:** Using a subset of the Truveta data we examine odds ratios (OR) of in-hospital remdesivir treatment and risk ratios (RR) of in-hospital length of stay between Black or African American (Black) to white patients. We adjusted for confounding factors such as age, sex, and comorbidity status.

**Results:** There were statically significant lower rates of remdesivir treatment and longer in-hospital lengths of stay comparing Black patients to white patients early in the pandemic (OR for treatment: 0.88, 95% confidence interval [CI]: 0.80, 0.96; RR for length of stay: 1.17, CI: 1.06, 1.21). Rates became close to parity between groups as the pandemic progressed.

**Conclusions:** While inpatient remdesivir treatment rates increased and length of stay decreased over the beginning course of the pandemic, there are still inequalities in patient care.

## Introduction

Inequalities in medical care in the United States (US) are widespread across socioeconomic (SES) and racial bounds^1^. The SARS-CoV-2 (COVID) pandemic has only intensified these inequalities^1^. Zelner et al.^2^ found a 5.5-fold increase in incidence and age-standardized mortality in Black or African American patients (Black) as compared to white patients, though their study was limited to the state of Michigan. In a systematic review of COVID outcomes, Mackey et al.^3^ showed that Black patients experienced higher rates of COVID infection and overall mortality, but not COVID-related mortality. However, no identified studies examined rates of COVID treatment or length of inpatient hospital stay (LoS). With ongoing calls for healthcare reform, racial equality, and the end of systemic racism, it is of key importance to identify areas where systematic inequalities have been bettered, where they still exist, and how they have changed over time.

Given the growing availability of effective COVID treatments over the course of the pandemic^4^, and the severity of unequal distribution of these treatments between racial groups, we aimed to assess the differences in treatment with remdesivir, as well as LoS between Black and white patients using one of the most complete, timely, and highest quality aggregation of electronic health records (EHR): the Truveta, Inc. data platform.

Remdesivir was the first antiviral given emergency use authorization by the US Food and Drug Administration (FDA) on May 1, 2020 for demonstration of effectiveness in reducing LoS in hospitalized patients^5,6^. It is an adenosine nucleoside triphosphate analog that interferes with viral RNA-dependent RNA polymerase^7^. It has been shown to significantly reduce COVID-related severity and duration of illness^8^. We focused on remdesivir because it is one of the most common COVID treatments with emergency use authorization being granted early in the pandemic and treatment rates are well represented in the Truveta dataset.

Electronic health record (EHR) data provide a comprehensive view of a patient’s journey through COVID prophylaxis, infection, symptomology, treatment, and ultimate outcome. Recent advances in data science, data security technology, and ethical and legal reviews of data storage, processing, and distribution have allowed unprecedented access to such data stores. For this study we aimed to examine the changes in delivery of COVID treatments and in-hospital LoS for COVID by race.

Results here show changes in differences between Black and white patient treatments with remdesivir and LoS with lower rates of treatment and longer lengths of stay for Black patients in the early stages of the pandemic, but becoming statistically indistinguishable during the Delta and Omicron waves after adjusting for a suite of potential comorbid confounders.

## Methods

### Study population

Our study included all inpatient COVID patients present in the de-identified Truveta electronic health records. Truveta provided the de-identified medical records used in this study on June 22, 2022. Truveta is a collective of healthcare systems which came together to aggregate EHR data with the purpose of enabling research. Currently this collective includes 24 members who provide patient care in over 20,000 clinics and 700 hospitals across 43 states. Updated data is provided daily to Truveta. Through syntactic normalization similar data fields from different systems are mapped to a common schema referred to as the Truveta Data Model (TDM). Once organized into common fields, the values are normalized to common ontologies such as ICD-10, SNOMED-CT, LOINC, RxNorm, CVX, etc., through semantic normalization. The normalization process employs an expert-led, artificial intelligence driven process to accomplish high-confidence modeling at scale. The data are then de-identified by expert determination under the HIPAA Privacy Rule. Once de-identified, the data are available for analysis in R or python using Truveta Studio. The Providence Health Care Institutional Review Board has declared this study not human research (STUDY2022000435). Our study period spanned from March 1, 2020 to March 1, 2022. Inclusion criteria included COVID-19 diagnosis, either from a lab result or an assigned diagnosis, during one of our treatment windows. Patients were excluded if their records were missing sex, and age information. For those patients who were missing encounter times and for those patients who received remdesivir, we excluded those patients missing drug administration times. The data provided by healthcare systems was comprised of patients from 40 US states.

We compared three remdesivir treatment windows corresponding to waves of the pandemic: “December 2020” from December 1, 2020 to February 28, 2021; “Delta” from June 1 to August 31, 2021; and “Omicron” from December 1 to February 28, 2021^9^. As remdesivir was given emergency authorization by the FDA on May 1, 2020, we included an additional wave – “wild type” from March 1 to May 31, 2020 in the LoS analysis.

Our main comparisons were between remdesivir treatment between Black patients and white patients as well as LoS in these two groups. Logistic regression with remdesivir treatment (yes/no) as the model outcome was used to estimate differences between racial groups adjusting for age and sex, as well as the following comorbidities: diabetes, hypertension, chronic kidney disease (CKD), liver disease, immunocompromised state, and cancer (ICD-10 and SNOMED codes are provided in the Supplementary Material). Poisson and Negative Binomial generalized linear regression models were fit to counts of days of LoS and compared by the Akaike information criterion (AIC)^10^ adjusting for the same covariates. We note that this is exploratory and does not account for censoring in days of LoS as a full survival analysis would due to privacy limitations with data re-identification issues. A Kolmogorov-Smirnov test was used to test for differences in ages between Black patients and white patients.

All analyses were conducted in R Version 4.1.3^11^, using packages `dplyr`^12^, `ggplot2`^13^, `gtools`^14^, `MASS`^15^, `xtable`^16^, `bit64`^17^, and `scales`^18^.

## Results

Of 556,134 patients who had a COVID diagnosis or lab test during one of the four pandemic periods of interest, 10.6% were inpatients, giving a final study population of 64,650 (Table 1). Females made up 51.2% of the inpatient sample, 14.6% Black, and 70.1% white. Due to the small proportion of Asian, American Indian or Alaska Native, Native Hawaiian or other Pacific Islander, or patients missing data on race, we focus only on Black and white patients in these results. Near 46% of the population was over 65 years of age. The distributions of ages was significantly shifted to younger ages for Black patients than for white patients (Kolmogorov-Smirnov test, D^+^ = 0.159, p *<* 0.001; Figure 1). Median age of hospitalizations for Black patients was 58.7 years (25%–75% quantile: 44.5 – 69.9) compared to 65.4 years (25%–75% quantile: 51.5 – 76.9) for white patients. Both groups were matched on percentage with other prevalent comorbidities as the epidemic progressed (Table 2).

**Figure 1.**
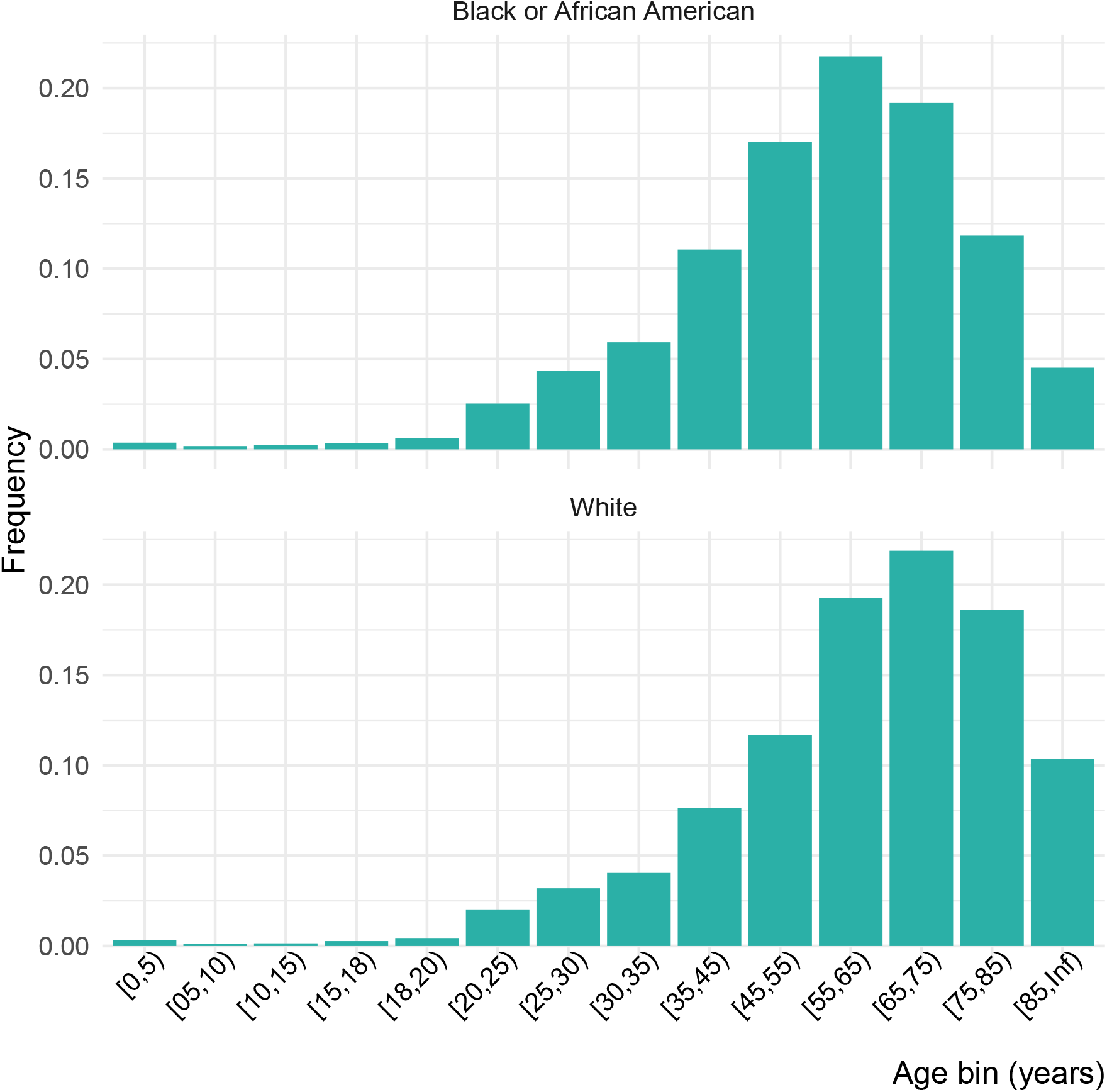
Distribution of ages by age bin, and race.

**Figure 2.**
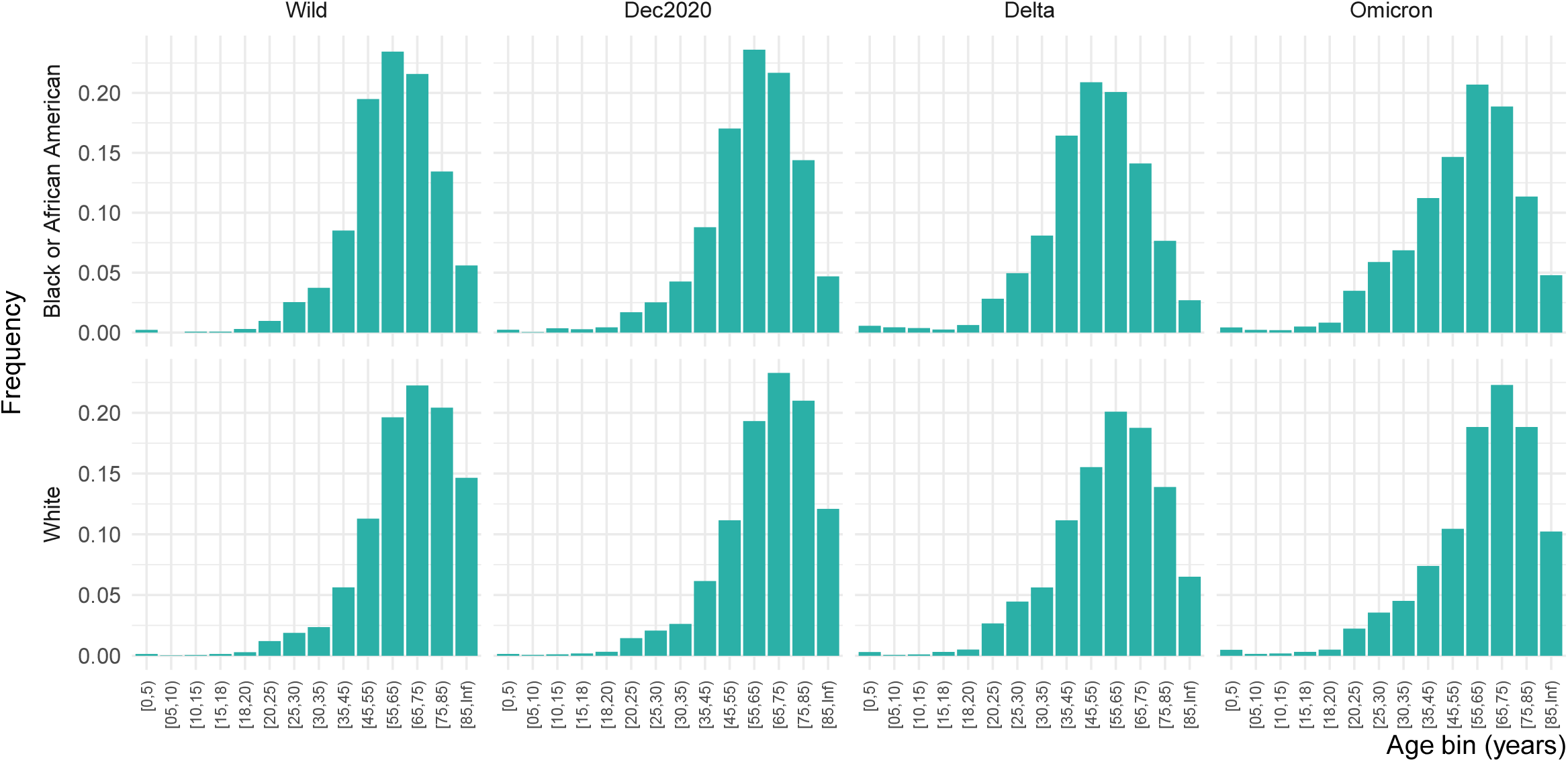
Distribution of ages by age bin, race, and pandemic wave.

**Table 1.** Demographic characteristics of the population affected by each wave of COVID-19 and their comorbidities (the age distributions are given in Figure 2).

|  | Wild<br>(N=5803) | Dec2020<br>(N=17824) | Delta<br>(N=12287) | Omicron<br>(N=28736) | Overall<br>(N=64650) |
| --- | --- | --- | --- | --- | --- |
| Sex |  |  |  |  |  |
| Female | 2840 (48.9%) | 8781 (49.3%) | 6355 (51.7%) | 15094 (52.5%) | 33070 (51.2%) |
| Male | 2963 (51.1%) | 9043 (50.7%) | 5932 (48.3%) | 13642 (47.5%) | 31580 (48.8%) |
| Race |  |  |  |  |  |
| American Indian or Alaska Native | 27 (0.5%) | 102 (0.6%) | 107 (0.9%) | 229 (0.8%) | 465 (0.7%) |
| Asian | 177 (3.1%) | 522 (2.9%) | 244 (2.0%) | 615 (2.1%) | 1558 (2.4%) |
| Black or African American | 1339 (23.1%) | 2537 (14.2%) | 1594 (13.0%) | 3991 (13.9%) | 9461 (14.6%) |
| Native Hawaiian or Other Pacific Islander | 18 (0.3%) | 64 (0.4%) | 68 (0.6%) | 112 (0.4%) | 262 (0.4%) |
| White | 3393 (58.5%) | 12553 (70.4%) | 8789 (71.5%) | 20602 (71.7%) | 45337 (70.1%) |
| Missing | 849 (14.6%) | 2046 (11.5%) | 1485 (12.1%) | 3187 (11.1%) | 7567 (11.7%) |
| Ethnicity |  |  |  |  |  |
| Hispanic or Latino | 889 (15.3%) | 2985 (16.7%) | 1994 (16.2%) | 3536 (12.3%) | 9404 (14.5%) |
| Not Hispanic or Latino | 4670 (80.5%) | 14350 (80.5%) | 9897 (80.5%) | 24250 (84.4%) | 53167 (82.2%) |
| Missing | 244 (4.2%) | 489 (2.7%) | 396 (3.2%) | 950 (3.3%) | 2079 (3.2%) |
| Age bracket |  |  |  |  |  |
| [0,5) | 12 (0.2%) | 42 (0.2%) | 55 (0.4%) | 209 (0.7%) | 318 (0.5%) |
| [05,10) | 3 (0.1%) | 18 (0.1%) | 25 (0.2%) | 63 (0.2%) | 109 (0.2%) |
| [10,15) | 3 (0.1%) | 30 (0.2%) | 30 (0.2%) | 68 (0.2%) | 131 (0.2%) |
| [15,18) | 7 (0.1%) | 44 (0.2%) | 47 (0.4%) | 124 (0.4%) | 222 (0.3%) |
| [18,20) | 22 (0.4%) | 69 (0.4%) | 82 (0.7%) | 186 (0.6%) | 359 (0.6%) |
| [20,25) | 79 (1.4%) | 316 (1.8%) | 379 (3.1%) | 825 (2.9%) | 1599 (2.5%) |
| [25,30) | 137 (2.4%) | 451 (2.5%) | 605 (4.9%) | 1296 (4.5%) | 2489 (3.9%) |
| [30,35) | 181 (3.1%) | 589 (3.3%) | 783 (6.4%) | 1588 (5.5%) | 3141 (4.9%) |
| [35,45) | 452 (7.8%) | 1340 (7.5%) | 1597 (13.0%) | 2508 (8.7%) | 5897 (9.1%) |
| [45,55) | 831 (14.3%) | 2272 (12.7%) | 2043 (16.6%) | 3259 (11.3%) | 8405 (13.0%) |
| [55,65) | 1218 (21.0%) | 3577 (20.1%) | 2403 (19.6%) | 5368 (18.7%) | 12566 (19.4%) |
| [65,75) | 1241 (21.4%) | 4023 (22.6%) | 2097 (17.1%) | 5952 (20.7%) | 13313 (20.6%) |
| [75,85) | 996 (17.2%) | 3292 (18.5%) | 1467 (11.9%) | 4769 (16.6%) | 10524 (16.3%) |
| [85,Inf) | 621 (10.7%) | 1761 (9.9%) | 674 (5.5%) | 2521 (8.8%) | 5577 (8.6%) |
| Treated with Remdesivir |  |  |  |  |  |
| Yes | 222 (3.8%) | 6948 (39.0%) | 4716 (38.4%) | 4833 (16.8%) | 16719 (25.9%) |
| No | 5581 (96.2%) | 10876 (61.0%) | 7571 (61.6%) | 23903 (83.2%) | 47931 (74.1%) |
| Cancer |  |  |  |  |  |
| Yes | 318 (5.5%) | 965 (5.4%) | 626 (5.1%) | 1870 (6.5%) | 3779 (5.8%) |
| No | 5485 (94.5%) | 16859 (94.6%) | 11661 (94.9%) | 26866 (93.5%) | 60871 (94.2%) |
| Chronic kidney disease (CKD) |  |  |  |  |  |
| Yes | 379 (6.5%) | 1432 (8.0%) | 738 (6.0%) | 2197 (7.6%) | 4746 (7.3%) |
| No | 5424 (93.5%) | 16392 (92.0%) | 11549 (94.0%) | 26539 (92.4%) | 59904 (92.7%) |
| Diabetes |  |  |  |  |  |
| Yes | 1033 (17.8%) | 3353 (18.8%) | 1931 (15.7%) | 4774 (16.6%) | 11091 (17.2%) |
| No | 4770 (82.2%) | 14471 (81.2%) | 10356 (84.3%) | 23962 (83.4%) | 53559 (82.8%) |
| Hypertension |  |  |  |  |  |
| Yes | 2113 (36.4%) | 6617 (37.1%) | 3941 (32.1%) | 10382 (36.1%) | 23053 (35.7%) |
| No | 3690 (63.6%) | 11207 (62.9%) | 8346 (67.9%) | 18354 (63.9%) | 41597 (64.3%) |
| Liver disease |  |  |  |  |  |
| Yes | 219 (3.8%) | 668 (3.7%) | 564 (4.6%) | 1557 (5.4%) | 3008 (4.7%) |
| No | 5584 (96.2%) | 17156 (96.3%) | 11723 (95.4%) | 27179 (94.6%) | 61642 (95.3%) |
| Immunocompromised |  |  |  |  |  |
| Yes | 405 (7.0%) | 1208 (6.8%) | 831 (6.8%) | 2468 (8.6%) | 4912 (7.6%) |
| No | 5398 (93.0%) | 16616 (93.2%) | 11456 (93.2%) | 26268 (91.4%) | 59738 (92.4%) |

**Table 2.** Percentages of each population (Black patients and white patients) for each comorbidity (the age distributions are given in Figure 2) by COVID wave.

| Race | Comorbidity | Wild | Dec. 2020 | Delta | Omicron | Overall |
| --- | --- | --- | --- | --- | --- | --- |
| Black or African American | Cancer | 52 (3.88%) | 115 (4.53%) | 56 (3.51%) | 195 (4.89%) | 418 (4.42%) |
| White | Cancer | 235 (6.93%) | 774 (6.17%) | 502 (5.71%) | 1486 (7.21%) | 2997 (6.61%) |
| Black or African American | CKD | 107 (7.99%) | 328 (12.93%) | 132 (8.28%) | 412 (10.32%) | 979 (10.35%) |
| White | CKD | 217 (6.4%) | 974 (7.76%) | 504 (5.73%) | 1565 (7.6%) | 3260 (7.19%) |
| Black or African American | Diabetes | 273 (20.39%) | 603 (23.77%) | 281 (17.63%) | 772 (19.34%) | 1929 (20.39%) |
| White | Diabetes | 566 (16.68%) | 2251 (17.93%) | 1340 (15.25%) | 3342 (16.22%) | 7499 (16.54%) |
| Black or African American | Hypertension | 541 (40.4%) | 1069 (42.14%) | 575 (36.07%) | 1574 (39.44%) | 3759 (39.73%) |
| White | Hypertension | 1283 (37.81%) | 4774 (38.03%) | 2887 (32.85%) | 7697 (37.36%) | 16641 (36.71%) |
| Black or African American | Immunocompromised | 79 (5.9%) | 158 (6.23%) | 96 (6.02%) | 313 (7.84%) | 646 (6.83%) |
| White | Immunocompromised | 285 (8.4%) | 936 (7.46%) | 636 (7.24%) | 1876 (9.11%) | 3733 (8.23%) |
| Black or African American | Liver disease | 34 (2.54%) | 74 (2.92%) | 65 (4.08%) | 161 (4.03%) | 334 (3.53%) |
| White | Liver disease | 138 (4.07%) | 474 (3.78%) | 405 (4.61%) | 1165 (5.65%) | 2182 (4.81%) |

Including adjustment for diabetes, hypertension, CKD, liver disease, immunocompromised state, and cancer, Black patients received remdesivir at a statistically significant 12% during the December 2020 wave (Table 3 and Figure 3). No statistical difference was seen for the Delta or Omicron waves, which were 9% lower and 7% lower odds than white patients, respectively.

**Table 3.**
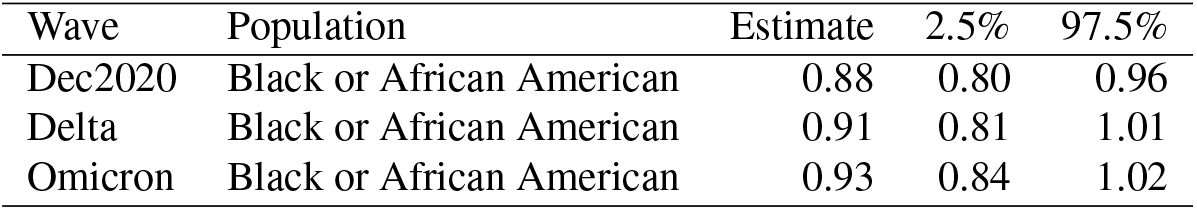
Odds Ratio and 95% confidence interval for remdesivir treatment while hospitalized for COVID comparing Black to white inpatients. Adjusted for age, sex, diabetes, hypertension, CKD, liver disease, immunocompromised state, and cancer.

| Wave | Population | Estimate | 2.5% | 97.5% |
| --- | --- | --- | --- | --- |
| Dec2020 | Black or African American | 0.88 | 0.80 | 0.96 |
| Delta | Black or African American | 0.91 | 0.81 | 1.01 |
| Omicron | Black or African American | 0.93 | 0.84 | 1.02 |

**Table 4.** Table shows the Risk Ratio of Length of Stay (LoS) for Black inpatients as compared to white inpatients. Adjusted for age, sex, diabetes, hypertension, CKD, liver disease, immunocompromised state, and cancer.

| Wave | Population | Estimate | 2.5% | 97.5% |
| --- | --- | --- | --- | --- |
| Wild | Black or African American | 1.13 | 1.06 | 1.21 |
| Dec2020 | Black or African American | 1.17 | 1.12 | 1.23 |
| Delta | Black or African American | 1.14 | 1.07 | 1.21 |
| Omicron | Black or African American | 1.03 | 0.99 | 1.07 |

**Figure 3.**
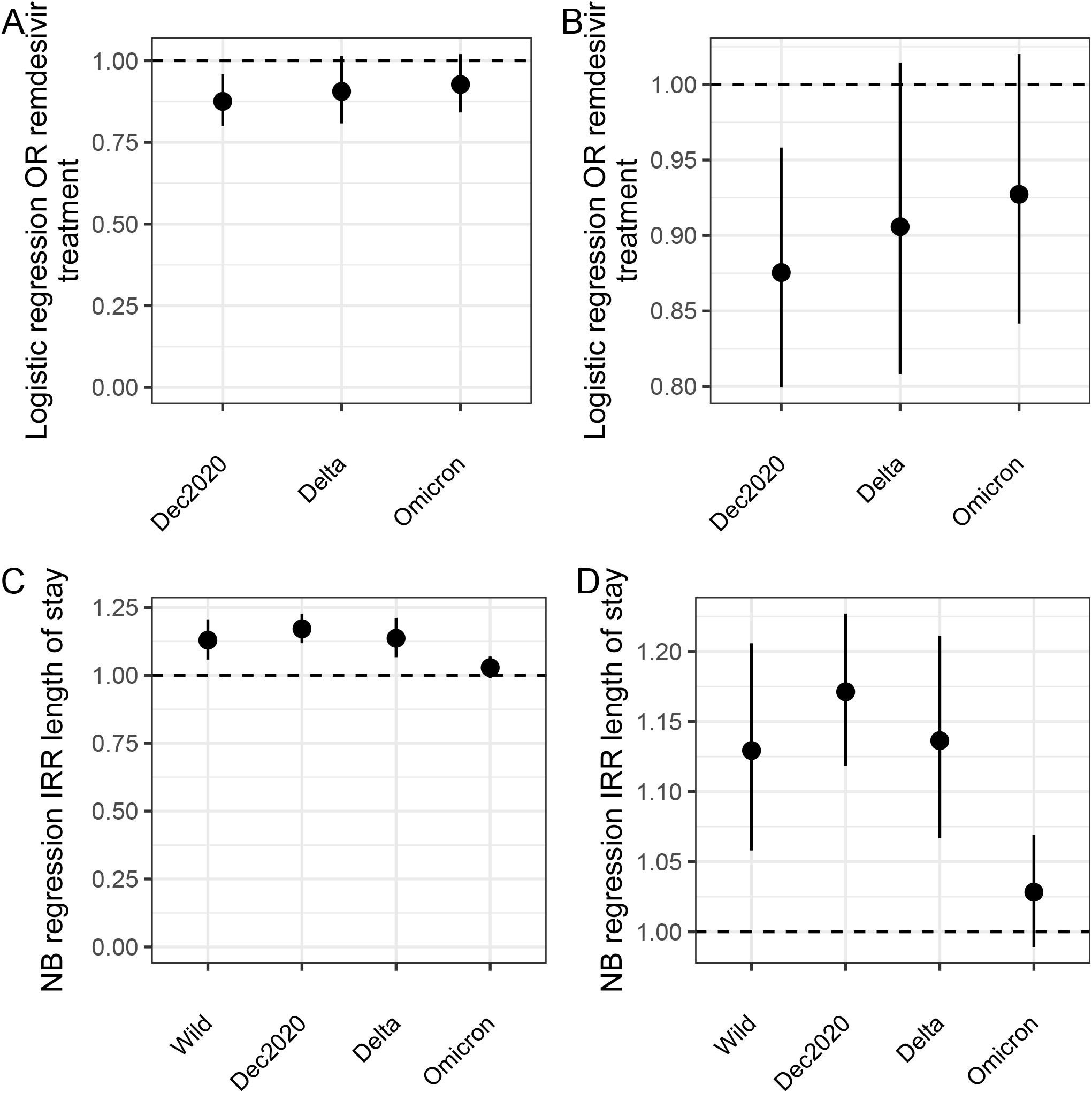
Logistic and NB regression results. **A & B** show the odds ratio for remdesivir treatment comparing Black to white inpatients. **C & D** show the risk ratio for LoS comparing Black to white inpatients. All analyses are adjusted for age, sex, diabetes, hypertension, CKD, liver disease, immunocompromised state, and cancer. Note the differences in y-axes between **A & C** and **B & D**.

Negative binomial models for days of LoS were preferred over Poisson models by AIC (Supplement). After adjustment for age, sex, diabetes, hypertension, CKD, liver disease, immunocompromised state, and cancer, Black inpatients had 5%, 16%, and 18% longer LoS for the wild type, December 2020, and Delta waves. There was no statistically significant difference for the Omicron wave (3%) (Table 3 and Figure 3).

## Discussion

Nearly all aspects of healthcare in our society have been affected disproportionally across SES, geography, and race or ethnic group. Unfortunately, the COVID pandemic has seen the same patterns^1^. Inequalities in access to healthcare, timeliness of treatment delivery, and treatment outcomes have been known for decades^19–24^. Our ongoing accumulation of knowledge on COVID outcomes, epidemiology, and treatments allows us to better situate disparities across racial group. Here we find that early in the pandemic there were broad differences in LoS and remdesivir treatments comparing Black patients and white patients with comorbidities. Hearteningly, our sample of over 60 thousand COVID inpatients, shows this gap may be closing and approaching parity between the two groups.

We examined the rates of comorbidities between Black and white patients in our sample and found statistical differences for diabetes hypertension, CKD, liver disease, immunocompromised state, and cancer between race or ethnic groups that attenuated over time (Table 1). In addition, we saw marked differences in the distribution of ages between groups who were hospitalized; Black patients were hospitalized at ages nearly 10 years younger than white patients, and an overall left shift in the age distribution for Black patients. This may indicate the earlier occurrence of the comorbidities in Black populations than white populations. This has been seen for diabetes^25^, hypertension^26,27^, CKD^28^, liver disease^29^, and cancer^30,31^. It could also be due to other behavioral and social factors (e.g. smoking status, or access to primary care). Future work should examine in more detail the drivers of this age discrepancy, and interventions implemented to reach younger Black individuals.

Geographic differences in susceptibility to COVID and treatment practice may also contribute to the disparities seen on a national scale. For example, one study showed that environments with high particulate matter were positively correlated with higher mortality in COVID^32^. COVID incidence peaked in different geographies at different times. Thus further study of differences across geography with respect to race or ethnic group is warranted. We did not explicitly incorporate geography into our analyses and leave that for future work.

Like all studies, ours is not without limitations. First, we compared only Black and white racial groups of patients for two reasons: 1) while the Truveta platform has representative data on race or ethnic group, there are still sample size limitations to race or ethnic groups other than Black and white; 2) there have been multiple studies examining various aspects in inequalities between Black and white groups, but few or none using data as large and comprehensive as in the platform, and many are from early in the pandemic^1,3,33–37^. Second, our estimates of differences in LoS between race or ethnic groups are calculated using Poisson and NB regression techniques which do not account for right- or left-censoring of LoS. Future work could use a survival analysis approach to adjust for censoring. Third, we studied waves of the pandemic corresponding to the dominant COVID strain at that time, and we do not include information on whether the patient had that dominant strain at the time. That said, the intent of the study was to characterize differences in remdesivir treatment (chosen because of remdesivir’s early emergency use authorization and for having a large enough sample size in our data) and not the specific infecting strain type. Fourth, we did not examine the potential effects of limitations in hospital capacity which may have exasperated disparities in treatment. Future work should address geographic differences in disparities and local hospital capacity. Fifth, the data cannot assess the time to diagnosis of COVID and the severity at admittance. Future studies should account for the full spectrum of social determinants of health that drive an individual to seek healthcare. Finally, the inclusion criteria for this study included individuals who had been hospitalized with a COVID infection, thus we cannot distinguish those admitted *for* COVID or those admitted for *something else* and COVID was an ancillary diagnosis.

Despite these limitations, ours is a large study of over 60 thousand patients hospitalized with COVID and shows differences in treatments and LoS between Black patients versus white patients. There are still many steps to be taken to achieve equity in healthcare for all. Policymakers should prioritize continuing to understand where disparities exist and achieving equity in access to healthcare and quality of care.

## Supporting information

Supplemental definitions and AIC comparisons

## Data Availability

Code necessary to generate all analyses and figures is included in a GitHub repository at

https://github.com/Truveta/althouse_et_al_covid_equity

## Institutional Review Board Approval

The Providence Health Care Institutional Review Board has declared this study not human research (STUDY2022000435).

## Data Availability Statement

Code necessary to generate all analyses and figures is included in a GitHub repository https://github.com/Truveta/althouse_*e*_*t*_*a*_*l*_*c*_*ovid*_*e*_*quity*.

