## Supplemental definitions and AIC comparisons for "COVID treatment and in-hospital length of stay inequalities between race in the US over time"

### Data element definitions

Below are the codes used for defining the COVID outcome as well as for the comorbidities.

**Remdesivir treatment:**
RxNorm: 2367757, 2367758, 2284718, 2284957, 2284958, 2284959, 2284960, 2395503, 2395499, 2395500, 2395502, 2395504

#### COVID Outcome

**COVID:**
ICD-10: B97.29, U07.1, Z86.16;
SNOMED CT: 840539006, 870588003, 870589006, 870590002, 870591003, 119731000000000, 119741000000000, 119751000000000, 119981000000000, 1240520000000000, 441590008, 840544004, 1017214008, 1119302008

#### Comorbidities

**Cancer:**
ICD-10: C00–C26, C40–C96

**CKD:**
D63.1, E08.22, E09.22, E13.22, I12, I12.0, I12.9, I13, I13.10, I13.11, I13.2, N18, N18.1, N18.2, N18.3, N18.4, N18.5, N18.6, N18.9, O10.211, O10.212, O10.213, O10.219, O10.22, O10.31, O10.32, O10.33, Q61.2, Q61.3, Q61.8, Z94.0

**Diabetes:**
ICD-10: E10, E11, E13

**Hypertension:**
ICD-10: I10, I11, I12, I13, I15

**Immunocompromised:**
ICD-10: B20, C00–C26, C40–C96, D00–D09, D37–D44, D80–D84, D89, Z94

### AIC comparisons of Poisson and Negative Binomial regressions

| Population | AIC Poisson | AIC NB | Delta AIC |
|---|---|---|---|
| Dec2020 | 123955.63 | 71840.52 | -52115.11 |
| Delta | 100568.24 | 51387.44 | -49180.80 |
| Omicron | 223433.61 | 125462.91 | -97970.70 |
